# Teaching an Old Dog New Tricks: Serum Troponin T as a Biomarker in Amyotrophic Lateral Sclerosis

**DOI:** 10.1101/2021.02.15.21251783

**Authors:** Sergio Castro-Gomez, Barbara Radermacher, Pawel Tacik, Sandra R. Mirandola, Michael T. Heneka, Patrick Weydt

## Abstract

Amyotrophic lateral sclerosis (ALS) is a devastating neurodegenerative disease characterized by progressive loss of upper and lower motor neurons. Diagnosis, management and therapeutic trials are hampered by a lack of informative biomarkers. Troponins (Tn) are components of skeletal and cardiac muscles. Acute elevation of cardiac isoforms of troponin I (cTnI) and T (cTnT) in serum indicates myocardial injury. Case reports suggested that serum levels of cTnT, but not cTnI are chronically elevated in ALS and other neuromuscular disorders.

Using standard clinical laboratory methodologies we studied serum troponin levels in a multicentric cross-sectional cohort of 75 ALS patients and sixty controls (DESCRIBE-ALS cohort) and in a real-world cohort of 179 consecutive patients from our ALS clinic at the University Hospital Bonn.

We found that serum cTnT, is elevated in >60% of ALS patients while cTnI is always normal. Serum cTnT levels increase over time and correlate with disease severity as measured with the revised ALS FRS score. There was no correlation with the phosphorylated neurofilament heavy chain (pNfH) levels in the cerebrospinal fluid. We propose that cTnT elevations in ALS are of non-cardiac origin and may serve as a proxy of lower motor neuron or skeletal muscle involvement. They potentially help to stratify patients according to lower motoneuron involvement. Further research will determine the biological origin of the cTnT elevation and its validity as a diagnostic and/or prognostic marker. Our finding also serves as a reminder to interpret cTnT with caution elevations in patients with neuromuscular diseases.

## Introduction

Amyotrophic lateral sclerosis (ALS), the most common form of adult-onset motoneuron diseases, remains a clinical diagnosis and is defined as the combination of progressive upper and lower motor neuron symptoms [1]. There is an urgent need for biomarkers that inform diagnosis, prognosis and the design of interventional trials [2].

Troponins (Tn) are essential structural and functional components of skeletal and cardiac muscle. Expression of cardiac troponin I (cTnI) and T (cTnT) is highly tissue specific and their elevation in serum is a sensitive and specific indicator of acute myocardial injury [3]. The interpretation of troponin serum levels is complicated by a range of chronic conditions associated with persistently elevated cardiac troponins. Serum troponin levels in ALS are rarely reported. A limited number of case reports and a case control study, however, suggest that cTnT is elevated in ALS even in the absence of evidence for cardiac damage [4–6]. This prompted us to speculate that serum cTnT levels might have an unexpected informative value for establishing the diagnosis and prognosis of ALS, especially if cTnI is measured simultaneously to control for potentially undetected myocardial injury.

Here we combined an observational cross-sectional biomarker study and real-world evidence from our ALS and memory clinics to test the hypothesis that serum cTnT elevation is a hallmark of ALS and reflects disease severity.

## Subjects and Methods

### Subject samples

We analysed clinical information and serum samples from two resources: (1) the DESCRIBE cohort is a multicentric observational study maintained by the German Center for Neurodegenerative Diseases (DZNE). It recruits patients with neurodegenerative conditions, including ALS, and age- and sex-matched healthy controls. All DESCRIBE participants provided written informed consent [7]. (2) The Department of Neurodegenerative Diseases and Gerontopsychiatry at Bonn University Hospital runs a large memory and ALS unit where cTnT and cTnI serum levels are part of the routine work-up.

### Laboratory markers

High-sensitivity Cardiac cTnT and cTnI measurements were performed in a fully accredited commercial laboratory (Labor Volkmann, Karlsruhe) or at the central laboratory of the University Hospital Bonn. All other parameters were routinely determined through the University Hospital Bonn central laboratory. The phosphorylated neurofilament heavy chain (pNfH) in CSF was measured at the University Medical Center of Ulm, Germany using standardised ELISA, as described previously [8]. Disease severity was assessed with the revised ALS-Fuctional Rating Scale (ALS-FRSr) [9].

S*tatistical analysis* was performed using IBM SPSS 21 and GraphPad Prism 8. χ^2^ and Fisher’s exact tests were used for categorical variables as appropriate. Significance values are shown in the tables uncorrected for multiple comparisons. Groups were compared for cTnT, cTnI levels and age using non-parametric Kruskal-Wallis test with pairwise comparisons adjusted for multiple comparisons controlling false discovery rate. Correlation analyses were performed using non-parametric Spearman correlations. Receiver operating characteristics (ROC) curves were calculated using non-parametric settings.

## Results

To ascertain the prevalence of troponin level elevations in ALS, based on a power analysis informed by the Mach *et al*. study [10], we obtained serum samples from three diagnosis groups in the DESCRIBE cohort: ALS (n=75), Alzheimer disease (AD) (n=29) and matched controls (n=30) (table 1).

**Table 1.**
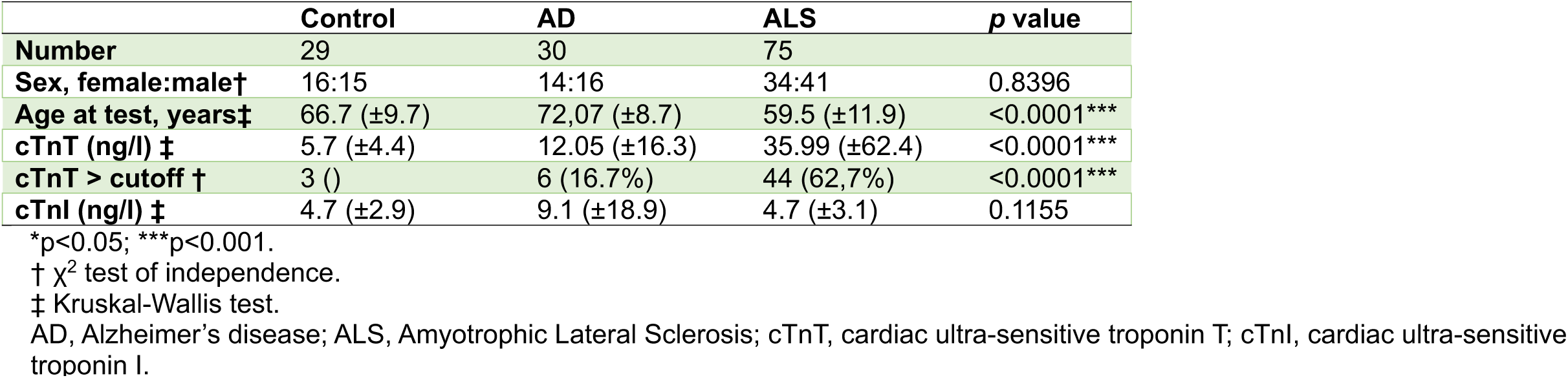
German center for neurodegenerative diseases-DZNE cohort.

The mean cTnT level in the ALS samples was 35.99 pg/ml (± 62.4 SD), significantly higher than in controls (5.7 pg/ml ± 4.4 SD) or AD (12.05 pg/ml ± 16.3) (Fig 1a). The mean cTnT level of the ALS cohort (35.99 pg/ml) was more than double the upper reference limit (14.0 ng/ml). In total 44 of the 75 ALS samples (62,7%) were above the upper reference limit. cTnI levels did not differ between the three conditions (Fig 1b) and were always within normal limits, except for two AD cases where however cTnT levels were also elevated (15.2/29.2 and 89.2/105.7 respectively).

**Figure 1.**
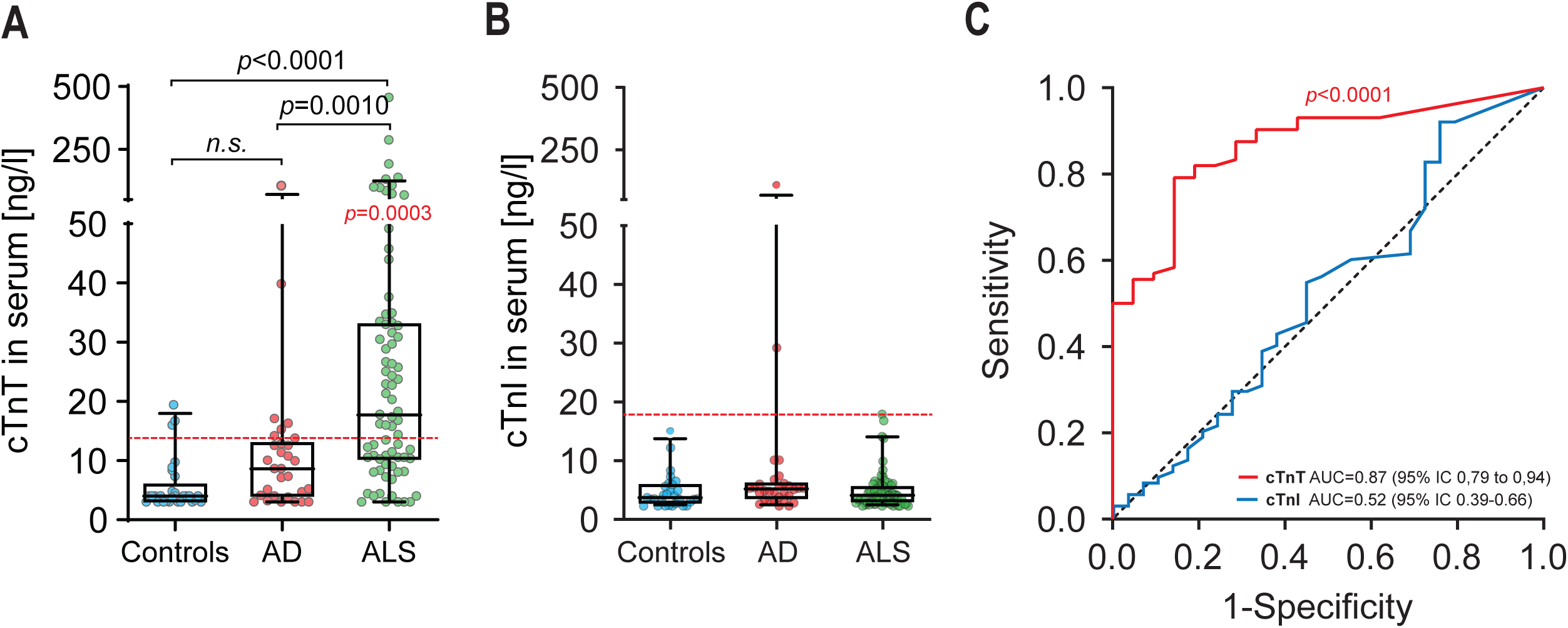
Cardiac troponin levels in serum from ALS patients. **(A)** cTnT levels in serum of control, AD and ALS patients (DZNE Cohort). The dashed horizontal red line is the conventional cut-off value (14 ng/l) for myocardial injury. Reported p (in black) are from Kruskal-Wallis tests on cTnT serum concentrations in comparisons between controls, AD and ALS patients. Reported p (in red) is from Wilcoxon Signed Rank Test on cTnT serum concentrations in comparison to the theoretical median of cut-off value of 14 ng/l for myocardial injury. Box plots represents the median and whiskers the 5-95th percentile. **(B)** cTnI levels in serum of control, AD and ALS patients. The dashed horizontal red line is the conventional cut-off value of 17 ng/l for myocardial injury. Box plots represents the median and whiskers the 5-95th percentile. **(C)** Receiver operating characteristic (ROC) curves illustrate cTnT (Red line) association with ALS diagnosis in comparison to cTnI levels in serum (blue line). cTnT = cardiac troponin T. cTnI = cardiac troponin T. AUC=area under the curve.

To determine the diagnostic accuracy of the two troponin tests for discriminating ALS from non-disease controls, we generated receiver operated characteristics (ROC) curves. The cTnT ROC curve had an area-under-the-curve (AUC) of 0.87. The cTnI ROC AUC was 0.52, and thus carried no information concerning the ALS diagnosis (Fig 1 C).

To better understand cTnT levels in the context of ALS, we interrogated real-world records from 117 consecutive patients that frequented our ALS unit from January 2019. In addition, 5 patients with benign fasciculation syndrome (BFS) were included as disease-mimics, and 38 AD and 19 Huntington’s disease (HD) patients as disease-control (Table 2). Because of a lag in implementing the test, cTnI levels were only available for a subset of the patients.

**Table 2.** Real-world cohort – University Hospital Bonn

| **Table 2 Real-world cohort – University Hospital Bonn** |  |  |  |  |  |  |  |
| --- | --- | --- | --- | --- | --- | --- | --- |
|  | **ALS** |  |  |  |  |  |  |
|  | **Spinal-onset** | **Bulbar-onset** | **PLS** | **BFS** | **AD** | **HD** | **p value** |
| **Number** | 85 | 24 | 8 | 5 | 38 | 19 |  |
| **Sex, female:male †** | 37:48 | 15:9 | 8:0 | 0:5 | 24:14 | 11:8 | 0.0025* |
| **Age, years ‡** | 64.4 (±11.6) | 64.2 (±8.5) | 59.9 (±8.2) | 41.5 (±9.0) | 73.8 (±8.4) | 52.4 (±10.6) | <0,0001*** |
| **Duration, years** | 3,1 (±2.8) | 2.5 (±1.8) | 7.4 (±7.9) | 3.5 (±4.2) | NA | NA | 0.3199 |
| **BMI (kg/m<sup>2</sup>) ‡** | 24.2 (±4.5) | 22.2 (±2.7) | 24.4 (±6.1) | 29.3 (±7.1) | ND | 27.9 (±6.7) | 0,1714 |
| **ALSFRS-R ‡** | 31.4 (±8.4) | 29.4 (±10.5) | 11.3 (±1.5) | 48 | ND | ND | <0,0001*** |
| **cTnT (ng/l) ‡** | 31.8 (±25.4) | 22.5 (±22.0) | 4.6 (±2.8) | 4.4 (±1.8) | 9.2 (±5.1) | 5.3 (±3.2) | <0,0001*** |
| **cTnT > cutoff †** | 64 (75,3%) | 11 (45,8%) | 0 (0%) | 0 (0%) | 7 (18.4%) | 0 (0%) | <0,0001*** |
| **cTnI (ng/l) ‡** | 6.6 (±4.3) | 5.5 (±2.2) | 5.4 (±2.7) | 5 (±1.3) | ND | 6.1 (±5.1) | 0,7577 |
| **pNfH (pg/ml) ‡** | 3058 (±2583) | 2330 (±1162) | 2541 (±1613) | 240.8 (±280.7) | ND | 355.4 (±295.5) | 0,0003*** |

\*p<0.05; \*\*\*p<0.001.
† $\chi^2$ test of independence.
‡ Kruskal-Wallis test.
AD, Alzheimer's disease; ALS, Amyotrophic Lateral Sclerosis; BFS, benign fasciculation syndrome; cTnT, cardiac ultra-sensitive troponin T; cTnI, cardiac ultra-sensitive troponin I; HD, Huntington's disease; PLS, Primary Lateral Sclerosis.

The mean cTnT level in the ALS cohort was 29,79 ng/ml (±24,89 SD), double the upper reference limit and similar to the DESCRIBE cohort (35.99 ng/ml). The mean cTnI level was 6,41 ng/ml (±3,95 SD), again similar to the DESCRIBE cohort (4.74 ng/ml).

The routine data collected in our clinic includes time and site of symptom-onset (bulbar vs. spine) and degree of upper motor neuron involvement, allowing us to classify three subgroups including the pure upper motor neuron variant primary laterals sclerosis (PLS) (Fig 2A). The highest absolute (131 ng/ml) and median (23.0 ng/ml) cTnT levels were detected in the spinal-onset group. The bulbar-onset grou (which usually also had limb involvement at the time of assessment), had a maximum cTnT level of 77.9 ng/ml and a median 13.6 ng/ml. AD patients presented with a cTnT mean value of 9.2 (±5.1 SD). All PLS, BFS and HD values were well below the upper limit of the reference range (<14 ng/ml).

**Figure 2.**
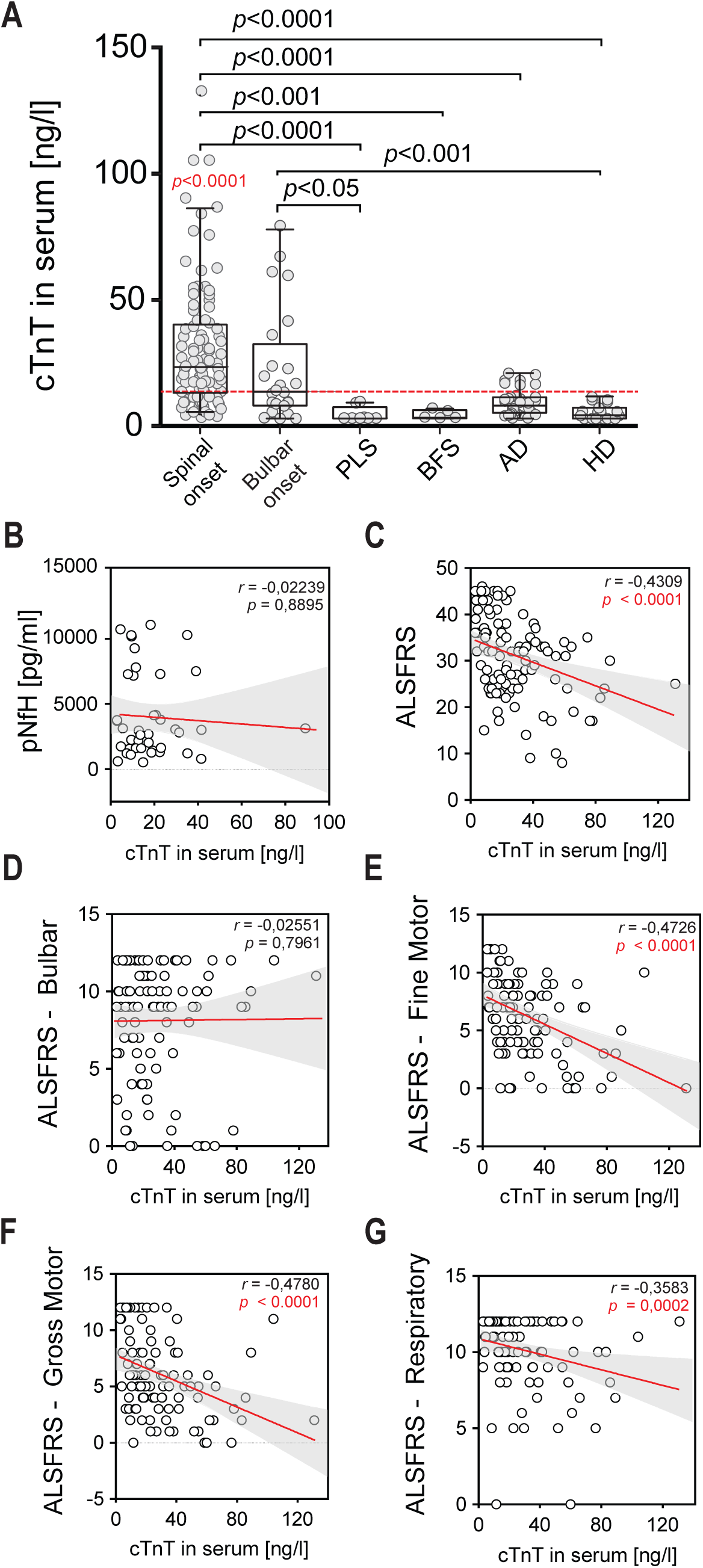
Real-life Cohort and correlation of cTnT in Serum with ALSFRS and sub-scores. **(A)** cTnT levels in serum of patients with ALS with limb-onset, bulbar-onset, PLS, BFS, AD and HD. Reported p (in black) are from Kruskal-Wallis tests on cTnT serum concentrations in comparisons between subgroups by Dunn’s multiple comparisons test. Reported p in red is from Wilcoxon Signed Rank Test on cTnT serum concentrations in comparison to the theoretical median of cut-off value of 14 ng/l for myocardial injury. The dashed horizontal red line is the established cut-off value of 14 ng/l for myocardial injury. Box plots represents the median and whiskers the 5-95th percentile. **B-G**. Correlation analyses were performed using non-parametric Spearman correlations (r). Curves were drawn by a linear regression model with an interaction term for cTnT in serum by **(B)** ALSFRS Global Score**(C)** ALSFRS Bulbar Score **(D)** ALSFRS Gross Motor Score **(E)** ALSFRS Gross Motor Score **(F)** ALSFRS Respiratory Score and **(F)** pNfH in CSF. Shaded areas represent 95% CIs. ALSFRS= Revised ALS functional rating scale, pNfH= phosphorylated neurofilament heavy chain, CSF= Cerebrospinal fluid.

We correlated the cTnT serum levels with total ALS-FRSr score and each of the subdomains, and -where available -with the pNfH levels in the CSF from the time of cTnT serum measurements (Fig. 2 B-G). cTnT showed a significant negative correlation with the total ALS-FRSr score. There was no correlation between cTnT and bulbar function, while the correlation with the fine and gross motor domain and with the respiratory domain was highly significant. There was no correlation between serum cTnT and CSF-pNfH.

In a subset of ALS patients (n=14) longitudinal data allowed us to examine the dynamics of cTnT and cTnI (Fig 3 B). Over time the individual cTnT levels in comparison to cTnI levels tended to increase. We then normalised the data by plotting the change of cTnT between the longest available interval (min. 30 days) as ΔcTn/day. The scatter of ΔTnT/day was significantly larger than ΔTnI/day (Fig 3 C), which did not show a significant change over time.

**Figure 3.**
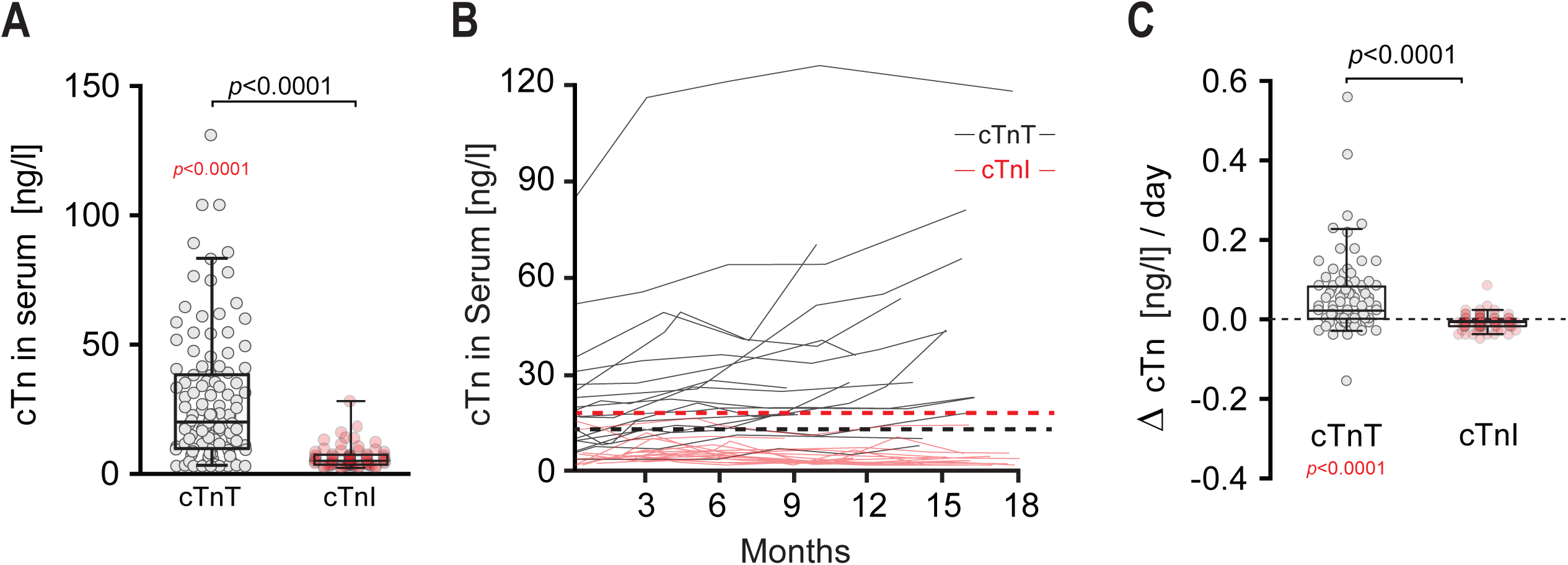
Correlation of cTnT Serum with ALSFRS and sub-scores. **(A)** cTnT in comparison to cTnI levels in serum of ALS patients (MND Clinic). Reported p (in black are from Mann-Whitney test on cTnT serum concentrations in comparisons to cTnI of ALS patients. Reported p in red is from Wilcoxon-Signed-Rank Test on cTnT serum concentrations in comparison to the theoretical median of cut-off value of 14 ng/l for myocardial injury. Box plots represents the median and whiskers the 5-95th percentile. **(B)** Spaghetti plot of cTn values versus time (months). Each black line represents longitudinal cTnT values of an individual patient. Each red line represents longitudinal cTnT values of an individual patient over time. Dash lines represent theoretical median of cut-off value of 14 ng/l (cTnT in black) and 17 ng/l (cTnI in red) for myocardial injury. **(C)** Change in time of cTnI und cTnT levels in serum of ALS patients when two measurements were available with a min. interval > 30 days. Reported p (in black) are from Mann-Whitney test on ΔcTnT serum concentrations in comparisons to cTnT. Reported p in red is from Wilcoxon Signed Rank Test on ΔcTnT serum concentrations in comparison to the theoretical median of no change. The dashed horizontal red line represents Δ=0. cTnT = cardiac troponin T. cTnI = cardiac troponin I. ΔcTn/day = (cTn first -cTn last)/days. Box plots represents the median and whiskers the 5-95th percentile.

## Discussion

This is, to the best of our knowledge, the first systematic report of a discordance between cTnT and cTnI levels in patients with ALS. We believe it has important and immediate implications for the laboratory work-up of patients with confirmed or suspected ALS.

The prior literature reveals a total of 11 cases with a similar cTnT vs cTnI discordance in four independent publications [4–6,11]. As case reports, none of these studies allow any conclusions on prevalence. A cohort study by Mach and colleagues found elevated cTnT in 68% of their population (n=40) vs. 5% in the case-control population [12]. This is in remarkably close agreement with our observations, but as Mach *et al*. did not measure cTnI, they had to speculate on the cardiac health of their subjects.

Our study offers the first opportunity to view the chronic cTnT elevation in ALS as a source of information rather than a confounding factor. The observation that cTnT levels correlate well with gross and fine motor deficits in the ALS-FRSr, but not with bulbar symptoms points to peripheral skeletal muscle as potential driver. Indeed, two independent research groups have put forward compelling arguments that degenerating or regenerating skeletal muscle is the likely source of the cTnT elevation in neuromuscular disorders, including ALS [4,5]. Of note, Bodor and colleagues report diaphragm muscle fibres expressing cardiac cTnT [13], while cardiac TnI is not detectable in any of the human skeletal muscle tested [12].

The lack of correlation between serum cTnT and pNfH in CSF is intriguing, as we may have two complementary biomarkers at hand: pNfH as a marker of neuraxonal damage reflecting upper motor neuron involvement and cTnT elevation as a proxy of neuromuscular, i.e. lower motoneuron affection. This is supported by the observation that patients with the upper motoneuron variant PLS had low normal serum cTnT. Clearly, research is necessary to determine the nature and origin of the elevated serum TnT levels in ALS patients. Also, our study reconfirms that in ALS, as in any neuromuscular disorder cTnT elevations need to interpreted with caution. We propose that simultaneously measuring cTnI is an efficient heuristic to rule out undetected myocardial injury when the clinical setting does not allow for an immediate and full cardiac work-up.

Finally, we note that troponins are emerging as therapeutic targets in ALS. Ongoing trials are looking at troponin C activators as potential disease modifiers [14]. We caution that troponin expression might be fundamentally altered in a large fraction of the ALS population and that this needs to be accounted for in trial design.

## Data Availability

The data in this manuscript can be made available upon reasonable request.

## Acknowledgments

The research was supported through unrestricted donations from our patients, especially Bruno Schmidt and the Initiative *Alle-Lieben-Schmidt e*.*V*. to PW.

## Author Contributions

All authors declare that they have participated in the design, execution, and analysis of the above study and that they have seen and approved the final version.

In detail: The study was conceived by SGC and PW. The study was designed by SCG, MTH and PW. Clinical data were obtained by SCG, BR, SM, PT, MTH and PW. SCG and PW analysed the data and PW drafted the manuscript with input from all co-authors.

